# Doubling time tells how effective Covid-19 prevention works

**DOI:** 10.1101/2020.03.26.20044644

**Authors:** Byung Mook Weon

## Abstract

Covid-19 (coronavirus disease 2019) is a rapidly spreading pandemic in many countries. The total confirmed cases of Covid-19 are exponentially increasing and many countries are fighting Covid-19 with all strategies. However, there is still lacking consensus for effective strategies. Here, I demonstrate the time dependence of the doubling time in the Covid-19 exponential growths. Tracking the time-dependent doubling time tells how well Covid-19 prevention works, giving an index for successful fighting.

---

The Cobid-19 has been spreading rapidly in many countries since it was first discovered in December 2019 [1, 2]. The total confirmed cases of Covid-19 for 12 countries are shown in **Fig. 1A**, retrieved from the Johns Hopkins University [3]. Each country has more than 4,500 cases as of March 23. The initial growths of the total cases for 12 countries exhibit the exponential growth dynamics, as in **Fig. 1B**. In general, the exponential growth of the total cases is represented by *N* = *N*_0_ exp(*γt*) where *N*_0_ and *N* are the total cases at time *t*_0_ and *t*, respectively, and *γ* is the growth rate. By definition, *N* = 2*N*_0_ needs the doubling time (*α*) that is expressed as *α* = ln(2)/γ through ln(*N* /*N*_0_) = ln(2) = *γt*. If the growth rate evolves with time, depending on the effective prevention strategies, then the doubling time changes with time. Thus, the time-dependent growth rate *γ*(*t*) causes the time-dependent doubling time *α*(*t*), defined as *α*(*t*) = *t* ln(2)/ ln(*N* /*N*_0_) through *γ*(*t*) = ln(*N* /*N*_0_)/*t*.

**FIG. 1:**
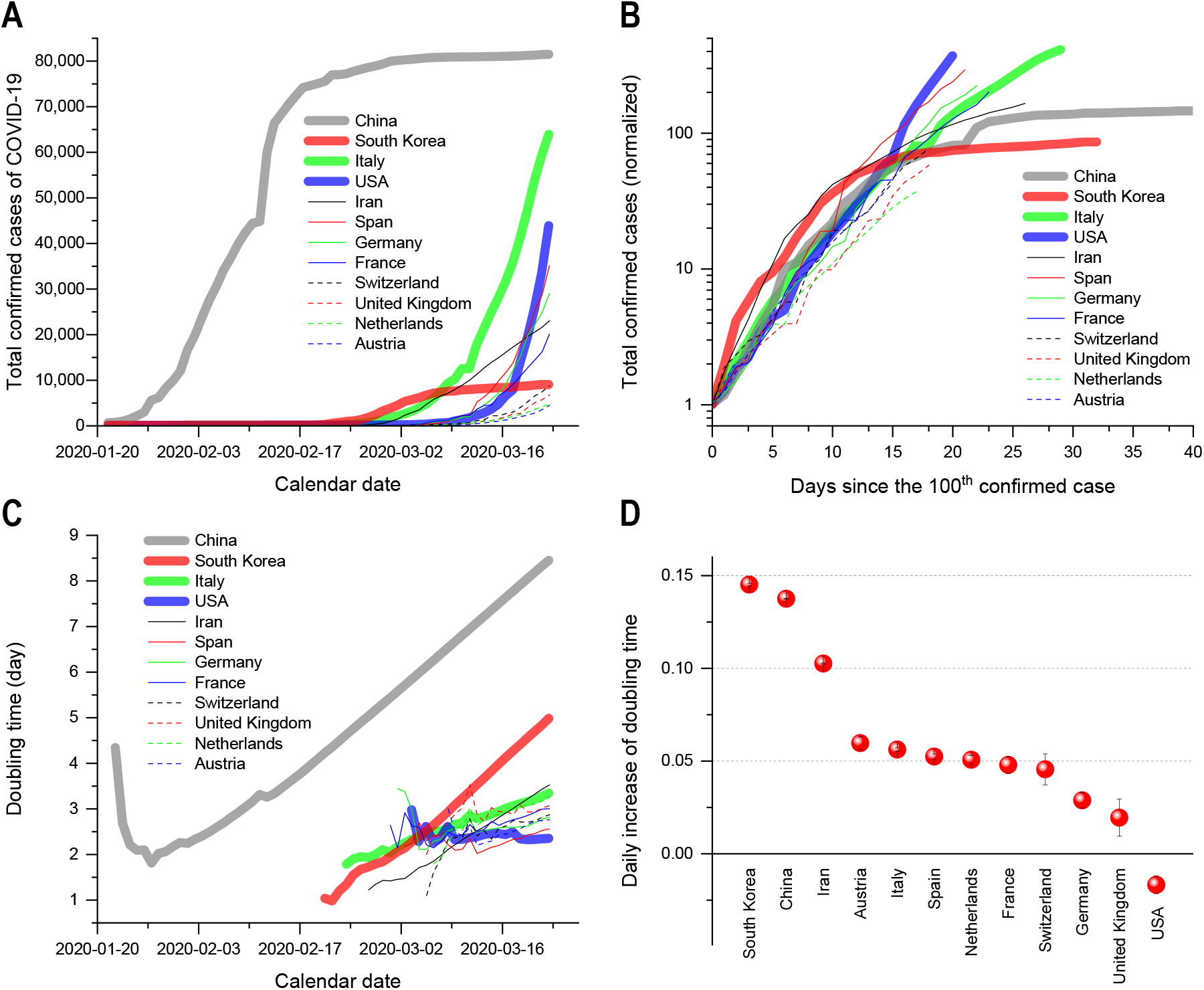
**A** The total confirmed cases of Covid-19 for 12 countries as of March 23, retrieved from the Johns Hopkins CSSE, show the drastic growths. **B** The total cases normalized by *N*_0_ show the exponential growths of the Covid-19 total cases (*N*). For reliable evaluations, *N*_0_ were chosen at *N* ≥ 100 (at or over the 100th case) and time was set to be 0 at *N*_0_ (see the same approach [2]; details of *N* and *N*_0_ in Appendix 1). **C** The doubling times taken from *α*(*t*) = *t* ln(2)/ ln(*N*/*N*_0_) demonstrate the time dependence of the doubling time (details of *α*(*t*) in Appendix 1). **D** The daily increases of the doubling times in 12 countries over the last 10 days (March 14 ∼ March 23) were estimated by the linear regression analyses with the software OriginPro (version 2019b). The error bars are the standard errors of the slopes (details of the regressions in Appendix 2). South Korea and China have the highest daily increases of the doubling times as ∼0.14 days per day, while India, Austria, Italy, Spain, Netherlands, France, and Switzerland have 0.10∼0.05 days per day, and finally Germany, UK, and USA have < 0.03 days per day.

The doubling time for 12 countries (**Fig. 1C**) was analyzed (**Appendix 1**). For China, the doubling time has declined until January 28 and then increased linearly since early February, due to the Wuhan containment since January 23. In South Korea, the doubling time has increased linearly without any initial decline. Since the early periods, South Korea has applied effective strategies, such as mass testing and patient tracking, to both symptomatic and asymptomatic patients [4]. The steady increases in the doubling times eventually lead to the flattened total case curves for China and South Korea [2]. In other countries, the increase of the doubling time was not sufficient to flatten the total case curves.

The daily increase of the doubling time for 12 countries over the last 10 days (March 14 ∼ March 23) (**Fig. 1D**) was estimated by linear regression (**Appendix 2**). In the last 10 days, South Korea and China have the highest daily increases of the doubling time (∼0.14 days per day), eventually achieving the doubling time > 4.0 days. India has ∼0.10 days per day and Austria, Italy, Spain, Netherlands, France, and Switzerland have ∼0.052 days per day (on average), not sufficient to get the flattening (the doubling time < 3.0 days). Finally, Germany, UK, and USA have < 0.03 days per day, implying little change in the doubling time. Little daily increase indicates that the initial exponential growth continues, implying no effective prevention strategies yet.

How effective preventive measures work [5] can be evaluated by tracking the doubling time. As demonstrated here, the time-dependent doubling time is real and informative. Successful fighting against Covid-19 would be achievable if the doubling time increases at the rate of ∼0.14 days per day, as seen in South Korea and China.

## Data Availability

All data are included in the supplementary files.

## Acknowledgments

I would like to thank Dr. Sun Do Hwang for useful discussion. The datasets of the total confirmed cases of Covid-19 were retrieved from the Johns Hopkins Center for Systems Science and Engineering Coronavirus Covid-19 Global Cases. This research was supported by Basic Science Research Program through the National Research Foundation of Korea (NRF) funded by the Ministry of Education (Grant No. 2019R1A6A1A03033215).

